# Analysis of the metabolic syndrome and its association with cART in HIV positive individuals receiving various cART regimens with review of literature

**DOI:** 10.1101/2022.03.08.22271951

**Authors:** Kavita S. Joshi, Udit U. Saraf, Rushabh Y. Gujarathi

## Abstract

**Context:** Many endocrine and metabolic disorders are seen in patients with HIV infection. Various comorbidities have been reported at a higher rate in HIV positive individuals, some at an earlier age. Since metabolic syndrome and its determinants are slowly developing, studies are needed in this regard.

**Aims:** The study aimed to analyze various parameters associated with the metabolic syndrome in HIV positive individuals and stratify subjects based on their treatment regimens, and present a brief comparison based on the same.

**Settings and Design:** A cross sectional study involving 155 participants was conducted at a tertiary care centre in Western India.

**Materials and Methods:** Detailed history and clinical examination was carried out. Routine investigations were done and parameters of interest to the study were then analysed based on AHA/NHLBI definitions.

**Statistical Analysis used:** Univariate analysis of all parameters. Multiple logistic regression for statistically significant parameters.

**Results and Conclusion:** Deranged HDL cholesterol was the most common component of the metabolic syndrome seen amongst all participants (*53*.*8%*) which was significantly higher in the treatment naïve group (*P = 0*.*001*). The difference between the incidence of metabolic syndrome between the ART naïve group and patients receiving ART was not significant. Males had a significantly higher prevalence of metabolic syndrome than females (*26*.*3%>12*.*4%, P = 0*.*026*). There was a significant difference in the incidence between the Zidovudine and Tenofovir treatment groups(*P=0*.*02*). Patients on the TLE (Tenofovir, Lamivudine, and Efavirenz) regimen had the lowest prevalence (*4*.*2%*) of metabolic syndrome.

## Introduction

According to the Sankalak Response - 2020, as published by the NACO (National AIDS Control Organisation), India is a low prevalence country in terms of HIV (Human Immunodeficiency Virus) with an estimated adult prevalence of 0.22% with 23.49 lakh PLHIV (People Living With HIV/AIDS) including an estimated 79,000 children living with HIV, constituting around 3.4% of the total PLHIV estimates.[^1^] Maharashtra continues to be a state with a prevalence of PLHIV being recorded above the national average at an estimated 0.36% with the largest numerical burden estimated to be 3.96 lakh. There were approximately 59,000 AIDS related deaths recorded in 2019 according to the same report. There has been an estimated 37% decline in the incidence of new HIV cases with an estimated 66% decline in AIDS related deaths between 2010-2019, much of it being attributed to a successfully running public health program in India (NACO), with the patients receiving combination antiretroviral therapy (cART).[^1^] HIV is now considered a chronic manageable disease.[^2^] The adult prevalence mentioned above accounts for the number of PLHIV in the age group of 15 to 49 years. These are the patients who are going to receive a long duration of cART, with treatment often initiated upon detection in accordance with the findings from various landmark clinical trials in the past decade.[^3,4^] Many endocrine and metabolic disorders are seen in patients with HIV infection, presumably due to their life expectancy now being almost the same as unaffected individuals, as reported by internationally conducted observational studies.[^5^] Studies have even reported an incidence of various chronic comorbidities at an earlier age than the general population in PLHIV, including but not limited to chronic liver disease, chronic kidney disease, diabetes mellitus and cardiovascular disease.[^6^] These may be a direct consequence of HIV infection, secondary to opportunistic infections or neoplasms, or related to medication side effects. Between 33 and 75% of patients with HIV infection receiving cART develop a syndrome often referred to as lipodystrophy, consisting of elevations in plasma triglycerides, total cholesterol, and apolipoprotein B, as well as hyperinsulinemia and hyperglycemia. Lipodystrophy is found to be more common amongst resource limited settings with a study from South India reporting a prevalence higher than 60%.[^7,8^] It has been suggested that the lipoatrophy changes are particularly severe in patients receiving the thymidine analogues Stavudine and Zidovudine with a switch from these drugs being associated with a qualitative improvement in findings related to lipoatrophy.[^9^] As per NACO, a rollout of Dolutegravir based regimens has been initiated and is expected to further reduce the AIDS disease burden.[^1^] However, a recent study has shown a significantly increased weight gain in patients receiving Dolutegravir with the tenofovir prodrug - Tenofovir Alafenamide (TAF), as compared to the standard treatment regimens.[^10^] This finding highlights the need for probing into metabolic syndrome related issues in PLHIV.

In the Indian general population, the estimated prevalence of metabolic syndrome was found to be 30%.[^11^] There is a dearth of studies focusing on metabolic syndrome amongst Indian PLHIV. An overall prevalence of around 19.8%, according to the Adult Treatment Panel III (ATP III) criteria, with an increased prevalence amongst patients receiving ART, was reported in a North Indian study.[^12^] There is a need to assess the metabolic syndrome amongst Indian PLHIV, with a focused analysis of various determinants related to both HIV and Metabolic Syndrome.

Since cART is a lifetime commitment, metabolic syndrome components tend to develop over time. It is thus envisaged that the findings of this study may point out the need for policy shifts concerning the testing for metabolic syndrome components amongst PLHIV. Moreover, knowing the extent of the problem will assist in taking appropriate precautionary measures in the management of HIV, with considerations being given to reduce the risk of developing metabolic syndrome. Hence, the present study was designed to be one of many aimed at addressing the aforementioned issues.

## Materials and Methods

### Study Type

It was a cross-sectional observational study carried out at a tertiary care centre in Western India.

### Study Population and Sample Size

HIV positive patients attending the Virology OPD of our hospital (Mumbai based tertiary care centre) were enrolled for the study. 155 Patients were recruited over a period of 12 months. Sample size was calculated using the OpenEpi software, as applicable to the number of PLHIV attended by the Virology OPD at our centre, at a 95% confidence interval.[^13^] The prevalence used for the same was the prevalence of metabolic syndrome in Indian PLHIV, which was reported to be 19.8%.[^12^]

### Selection of Participants

The inclusion criteria were patients aged more than 18 years, divided into 2 groups-Patients who were Combination Anti-Retroviral Therapy (cART) naïve and on cART for at least 6 weeks. Pregnant patients were excluded. Patients with a history of diabetes mellitus, hypertension, hyperlipidaemia, and patients on corticosteroids were also excluded. Study subjects were then divided into 3 groups

1. patients on Zidovudine based cART
2. patients on Tenofovir based cART
3. cART naïve

### Methods of Measurement

A detailed history was taken, and a clinical examination was done and recorded. The following measurements were recorded -Hip circumference, Waist circumference, Height, Weight. Fasting blood samples were collected from all study subjects. The following investigations were done. - Fasting and post-lunch blood sugar levels, Complete Lipid profile, CD4 count. It was an observational study, all investigations were done according to the standard treatment protocol with no additional investigations being added solely for the purpose of this study.

### Operational Definitions

The data for various parameters of metabolic syndrome was analysed on the basis of the normal limits as described by the American Heart Association/National Heart, Lung, and Blood Institute Scientific Statement for Asian populations which had slight modifications as compared to the previously used ATP III criteria.[^14^]

### Ethical Approval

Procedural approval was obtained from the regional Institutional Ethics Committee. Subjects were recruited in the study after obtaining written informed consent.

## Data Analysis/Statistics

Descriptive statistical analysis of data was done, and Chi-square and Fisher’s exact test were used for univariate analysis of the parameters being studied. Differences in the incidence of various outcomes amongst the groups being studied are mentioned as being statistically significant or insignificant based on the Chi-Square test. Multiple logistic regression was used for statistically significant parameters.

## Results

155 patients were enrolled based on inclusion and exclusion criteria. Out of these 155 patients, ten patients were later excluded, two patients were not compliant on cART, two patients had frequent changes of cART regimen, four patients were on Protease Inhibitor based regimens and two patients had been started on steroids before investigations. Hence these ten cases were excluded from further analysis. The remaining 145 patients were recruited, and their data was analyzed. 72 male and 73 female patients were enrolled. The difference in the sex distribution of patients on Zidovudine (Males>Females) and Tenofovir (Males<Females) can be attributed to the prevalence of anemia amongst Indian females. The mean age of the patients was found to be 40.1 ± 17.5 years (18-62 years). 75% of all patients were in the age group of 31-50 years.

The majority of patients in both groups had CD4 counts of below 200 at the at the initiation of cART. 56.25% of Zidovudine (*N = 48*) and 76.6% of Tenofovir (*N = 36*). Only 5 patients in the Zidovudine group and 2 patients in the Tenofovir group had CD4 counts of more than 350 at cART initiation. The mean initial CD4 count of patients in the Zidovudine group was 203.7 (*Standard Deviation = 100*.*8*) and in the Tenofovir group was 155 (*Standard Deviation = 100*.*9*). The mean duration of patients on stable ART was 229 weeks for the Zidovudine group and 85 weeks for the Tenofovir group. This difference was found to be statistically significant (*P = 0*.*0001*) and should be considered as a possible confounding factor while interpretation of the study findings.

High Blood Pressure (BP of above 130/85 mm Hg) was more common in the Zidovudine group (*N = 15, 31*.*25%*). The overall prevalence of elevated blood pressure was found to be 19.3% (*N = 28*). High blood pressure was more prevalent in patients on cART (*N = 23, 24*.*2%*) than those who were ART naïve. This difference was statistically significant (*P = 0*.*03*). Both, the Tenofovir (N=8, 17%) and the ART naïve (*N = 5, 10%*) groups had an overall lower prevalence of elevated blood pressure than the Zidovudine group (*N = 15, 31*.*25%)*. However, this difference was not statistically significant (*P = 0*.*25*). Patients on Zidovudine (*N = 15, 31*.*25%*) had higher BP than ART naïve patients (*N = 5, 10%*) with a statistically significant difference present between these two groups in particular (*P = 0*.*009*).

The overall prevalence of high waist circumference (≥ 90 cm in males, ≥ 80 cm in females) was 26.9% (*N = 39*). The difference between the incidences of high waist circumference was not significant between the different groups.

Deranged serum high-density lipoprotein (HDL) cholesterol (<40 mg/dL in males, <50 mg/dL in females) was the most common component of metabolic syndrome seen amongst all study subjects (*N = 78, 53*.*8%*). 88% ART-naive patients had deranged HDL cholesterol levels (*N = 44, 88%; P = 0*.*001*). This was significantly higher than both, the Zidovudine (*N = 15, 31*.*25%; P = 0*.*001*) and Tenofovir (*N = 19, 40*.*4%, P = 0*.*001*) groups.

31% patients (*N = 45*) had deranged serum triglyceride levels (≥150 mg/dL). The highest prevalence was found in the Zidovudine group (*N = 21, 43*.*75%*), which was significantly higher than the Tenofovir group (*N = 9, 19*.*1%; P = 0*.*008*).

Fasting blood sugar was deranged (≥ 100 mg/dL) in 14.5% of patients (*N = 21*). The Zidovudine group had the maximum number of patients with deranged FBS (*N = 10, 47*.*6%*). The Tenofovir group was found to have the lowest prevalence of deranged FBS (*N = 4, 8*.*5%*). However, this difference was not statistically significant.

A large number of patients satisfied 1 criterion of metabolic syndrome, both in the Zidovudine (*N = 13*) and Tenofovir groups (*N = 18*). This was followed by several patients not satisfying any criteria (*N = 13 for Zidovudine and N = 15 for Tenofovir*). Only 4 patients in the ART naive group did not satisfy any criteria for metabolic syndrome. This was due to the high prevalence of deranged HDL cholesterol (*88%*) in these patients. There was no patient in any group satisfying all the 5 criteria for the metabolic syndrome.

**Figure 1:**
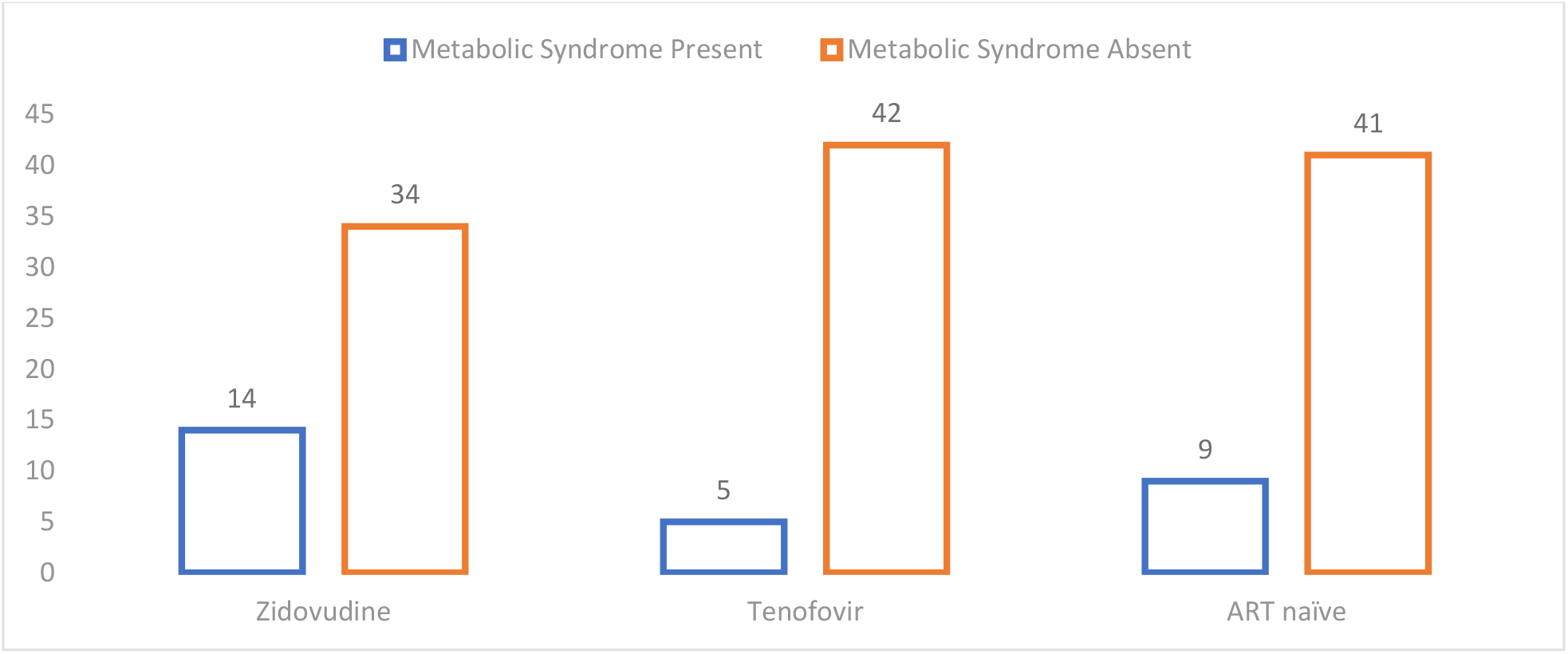
Prevalence of metabolic syndrome (≥3 criteria) in study population (n=145)

In the entire study population, 28 patients (19.3%) were diagnosed with metabolic syndrome as per the aforementioned criteria. The Zidovudine group had the highest prevalence of metabolic syndrome (N = 14, 29.2%) while the lowest was in the Tenofovir group (N = 5, 10.6%). This difference in the incidence of the metabolic syndrome between the two groups was found to be statistically significant (P = 0.02). 9 patients in the ART naive group were found to have metabolic syndrome as compared to 19 patients on ART. However, this difference was not found to be statistically significant (P = 0.48). 12.3% (N = 9) of female patients were found to have metabolic syndrome. Out of 9 female patients satisfying the criteria, only 1 was from the Tenofovir group, the group with the highest number of females. 19 (26.4%) males were found to have metabolic syndrome. The highest prevalence was in the Zidovudine group (N=11, 34.3%), followed by Tenofovir (N= 4, 23.5%) and ART naïve (N = 4, 17.4%) groups. The difference between the groups was not found to be statistically significant (P = 0.08). 26.4% of males (N = 19) and 12.3% of females (N = 9) were found to have metabolic syndrome. This increased prevalence of metabolic syndrome amongst males in the entire study population was statistically significant (P = 0.026). The ART naïve group did not show a significant difference in the prevalence between the two sexes (P = 0.07)

Patients on the TLE (Tenofovir; Lamivudine; Efavirenz) regimen had the lowest prevalence of metabolic syndrome (*N=1, 4*.*2%*). The difference in this incidence as compared to patients on the TLN regimen (Tenofovir; Lamivudine; Nevirapine) (*N = 4, 21%*) was not found to be statistically significant (*P = 0*.*16*). Patients on the ZLE regimen (Zidovudine; Lamivudine; Efavirenz) (*N= 4, 44*.*4%*) had a higher prevalence of metabolic syndrome than patients on the ZLN regimen (Zidovudine; Lamivudine; Nevirapine) (*N = 10, 25*.*6%*), although the difference was not significant (*P = 0*.*23*). Patients on the TLE regimen had a significantly lower prevalence of metabolic syndrome than both the ZLN (*P = 0*.*02*) and the ZLE (*P = 0*.*01*) regimens. Using multiple logistic regression considering these parameters, no significant difference was found between the Efavirenz and Nevirapine groups (*P = 0*.*08*). Other components of the metabolic syndrome had insignificant differences as listed in Table 3. Other differences between the groups were not significant statistically hence not mentioned.

**Table 1:** Demographic parameters in the three study groups

| Parameters |  | Zidovudine<br>Group | Tenofovir<br>Group | cART naïve<br>Group |
| --- | --- | --- | --- | --- |
| Gender | Male | 32 (66.6%) | 17 (36.1%) | 23 (46%) |
|  | Female | 16 (33.3%) | 30 (63.9%) | 27 (54%) |
| Age (years) | <30 | 3 (6.25%) | 11 (23.4%) | 3 (6%) |
|  | 31-40 | 19 (39.6%) | 16 (34%) | 24 (48%) |
|  | 41-50 | 19 (39.6%) | 12 (25.5%) | 20 (40%) |
|  | >50 | 7 (14.6%) | 8 (17%) | 3 (6%) |

**Table 2:**
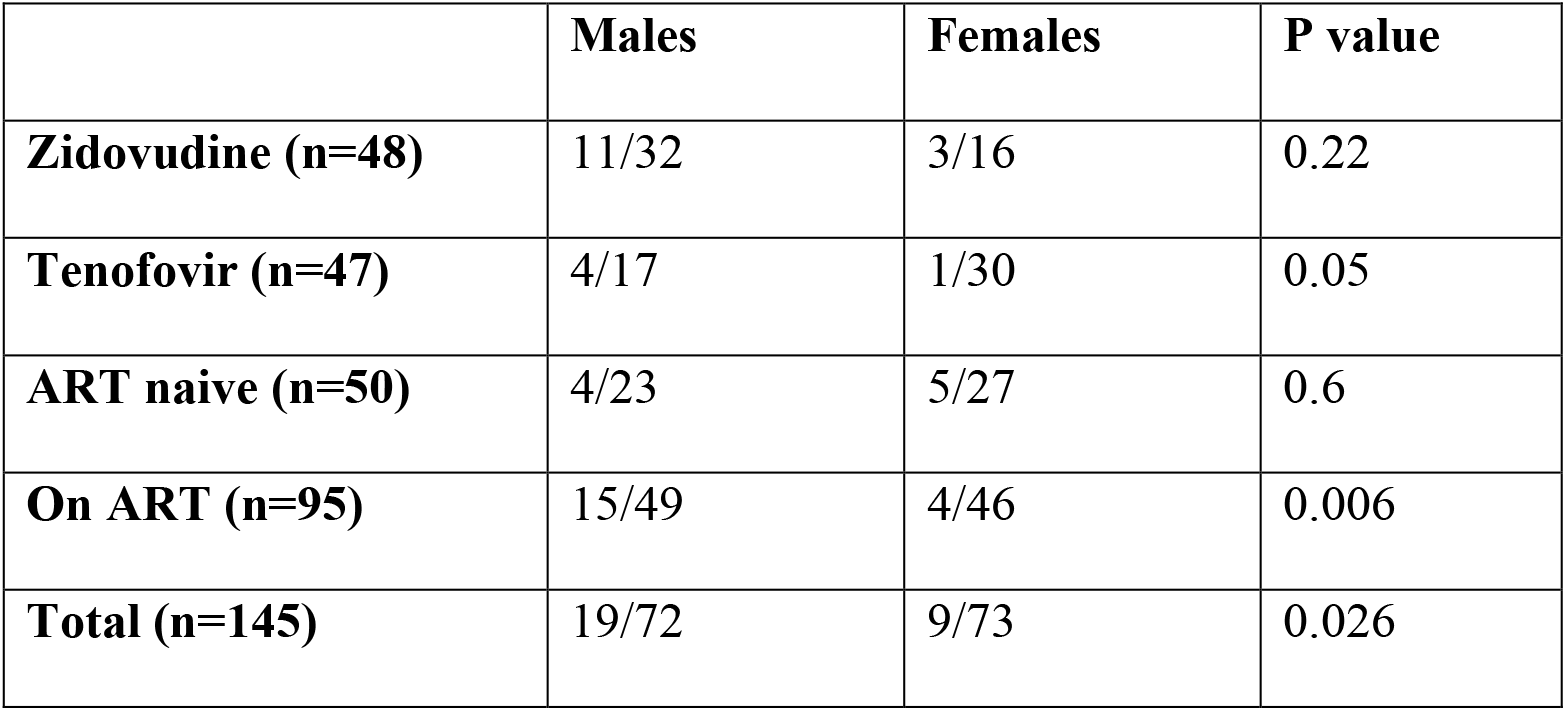
Comparison of metabolic syndrome based on Gender.

**Table 3:**
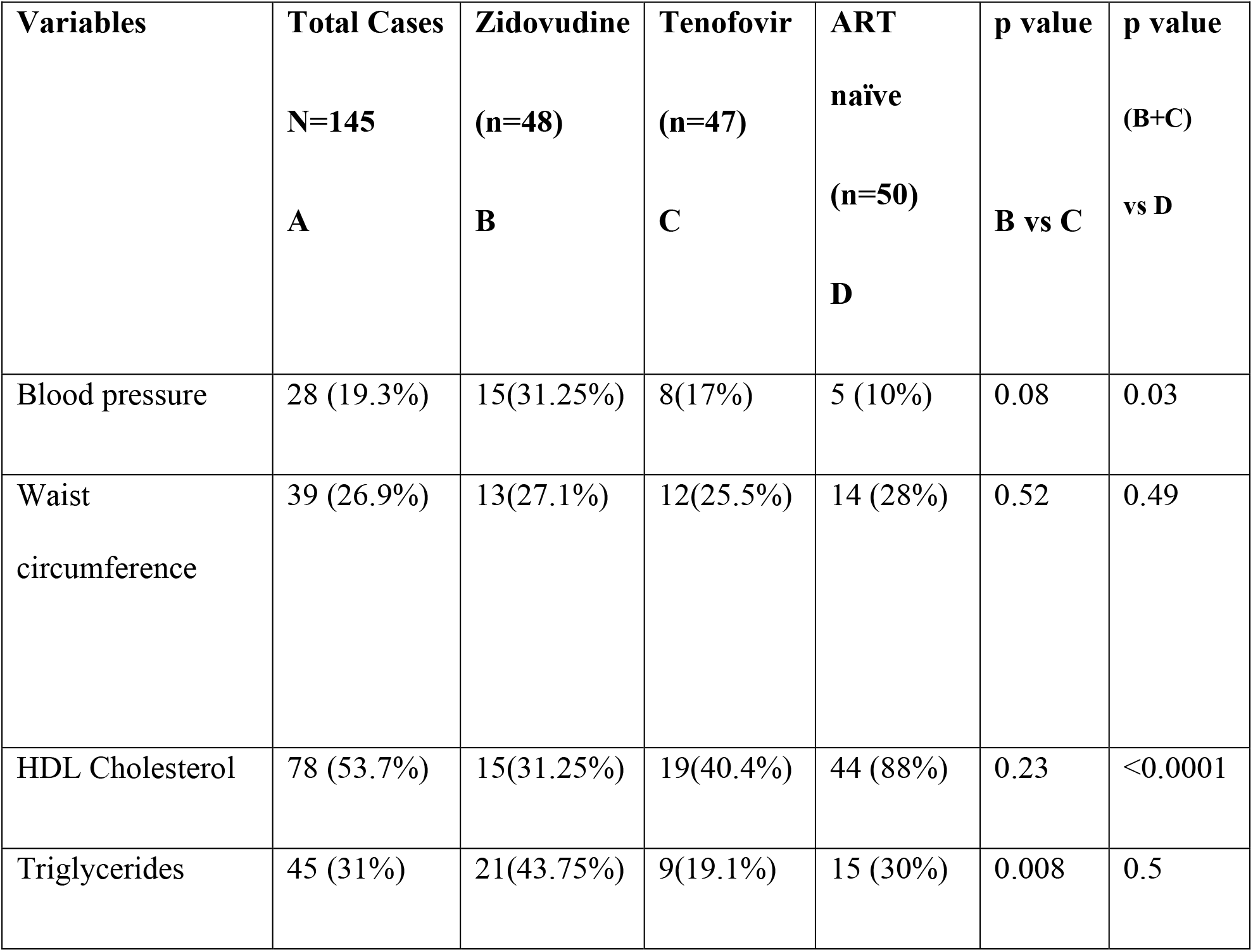
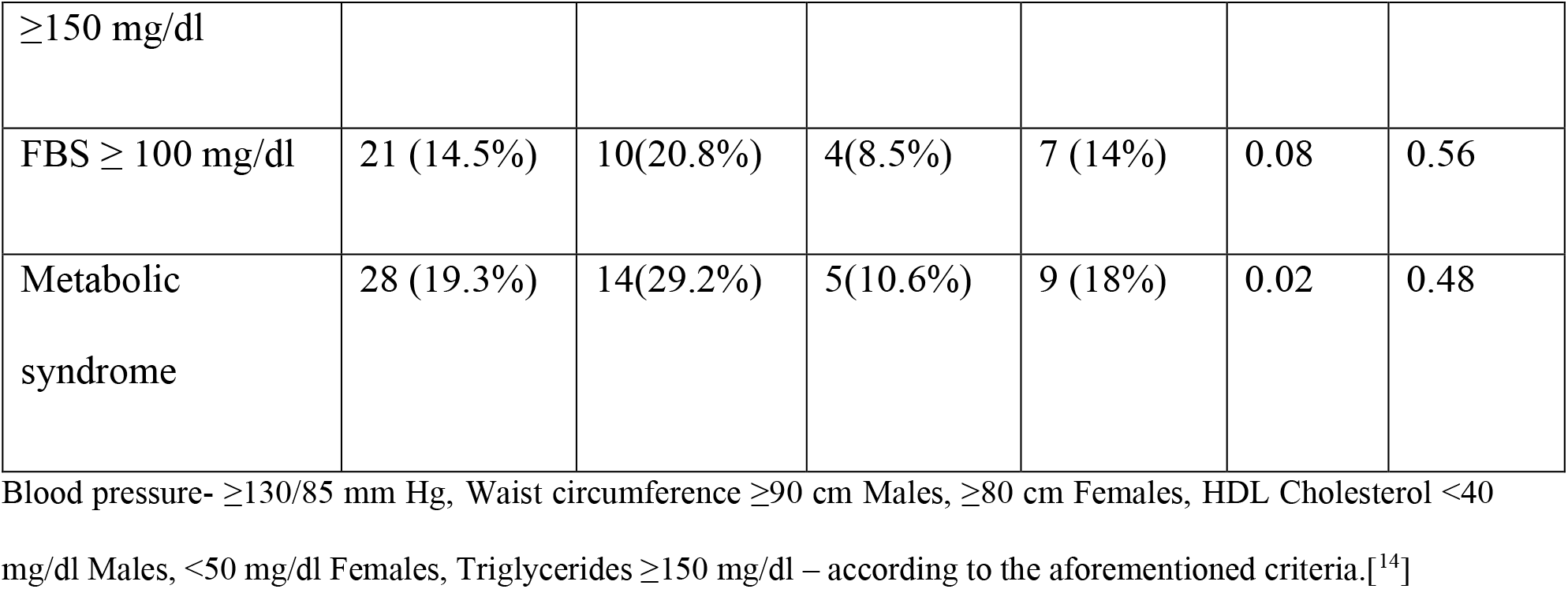
Comparison of metabolic syndrome in patients on Zidovudine vs Tenofovir:

**Table 4:** Comparison of Metabolic syndrome between studies.[^16,20,21,23^]

| Deranged Variables | Present study (n=145) | Bajaj et al. [20] (n=70) | Berhane et al. [21] (n=313) | Jerico et al. [16] (n=710) | Gazzaruso et al. [23] (n=533) |
| --- | --- | --- | --- | --- | --- |
| **Blood pressure** | 28 (19.3%) | 9 (12.85%) | 110 (35.1%) | 187<br>(26.3%) | 234 (42.3%) |
| **Waist circumference** | 39 (26.9%) | 0 | 19 (6%) | 89<br>(12.5%) | 209 (37.8%) |
| **HDL Cholesterol** | 78 (53.7%) | 34 (48.6%) | 149 (47.6%) | 253<br>(35.6%) | 290 (52.4%) |
| **Triglycerides** | 45 (31%) | 30 (42.85%) | 122 (39%) | 306<br>(43.1%) | 328 (59.3%) |
| **FBS (Fasting Blood Sugar)** | 21 (14.5%) | 20 (28.57%) | 78 (24.9%) | 82<br>(11.5%) | 133 (24%) |
| **Metabolic syndrome** | 28 (19.3%) | 14 (20%) | 66 (21.1%) | 121 (17%) | 251 (45.4%) |

**Table 5:**
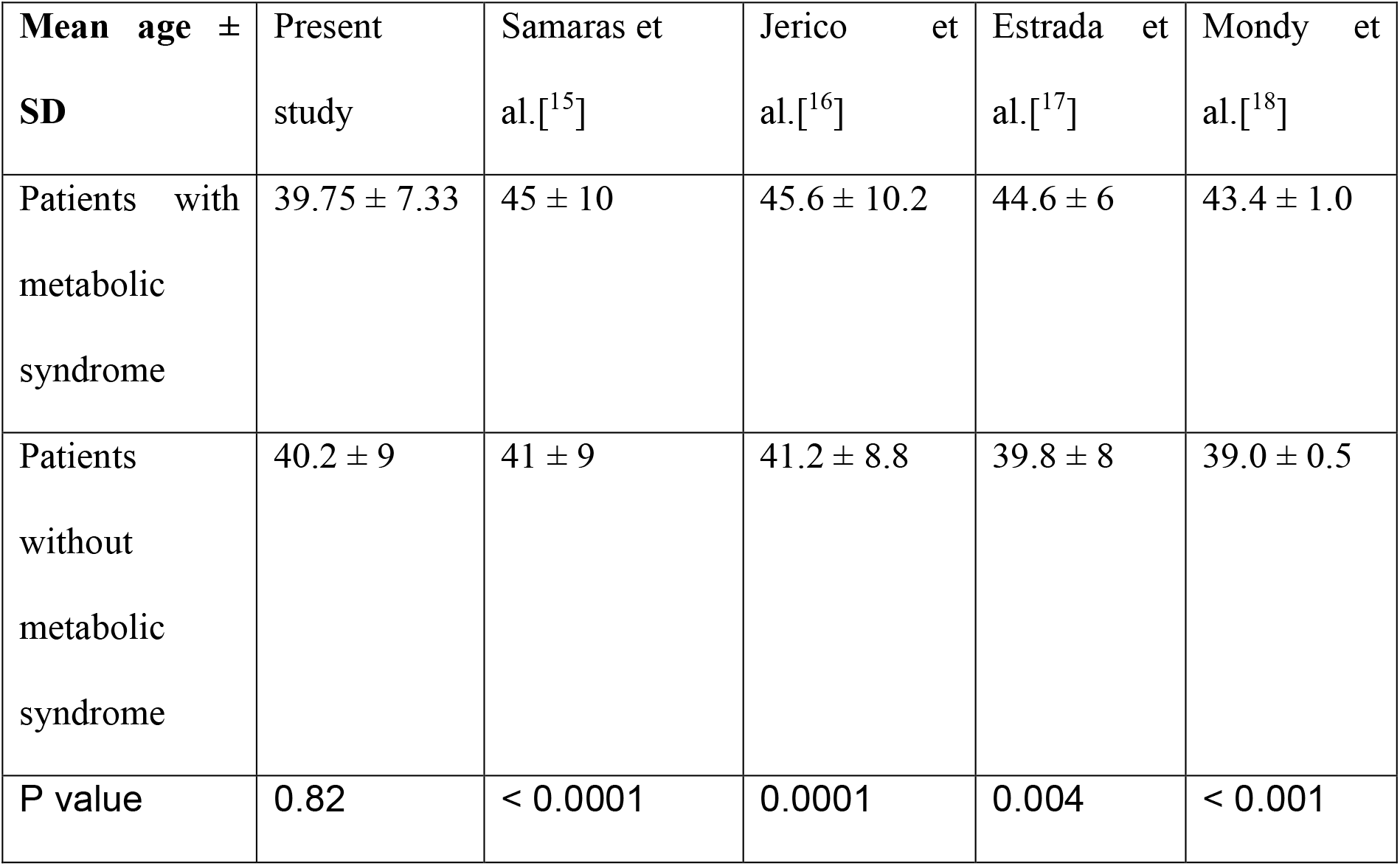
Age difference in metabolic syndrome between various studies.

## Discussion

The mean age of the patients in this study was found to be 40.1 ± 17.5 years (18-62 years). In the studies by Samaras et al., Jerico et al., Estrada et al. and Mondy et al., a higher age was significantly associated with an increased incidence of metabolic syndrome.[^15,16,17,18^] However, in the present study, there was no significant correlation between age and metabolic syndrome. A lack of statistical power through the current sample size may be responsible for this non-concordant finding. Alternatively, it may also mean that metabolic syndrome in our population occurs at a younger age than in populations around the world, especially amongst HIV affected individuals.

In the present study, an almost equal number of males and females were enrolled with the intention of eliminating confounding based on gender. This was comparable to a study by Jantarapakde et al.[^19^] However, other studies such as by Jerico et al and Bajaj et al had a higher number of male subjects (72% and 71.4% respectively).[^16,20^] The study by Berhane et al was the only study with more female patients (65.2%).[^21^] 73.9% of the patients were males in the study by Alvarez et al.[^22^] In the present study, males had significantly higher prevalence of metabolic syndrome than females. This contrasted with the study by Alvarez et al where females had significantly higher prevalence of metabolic syndrome than males.[^22^] In studies by Jerico et al, Mondy et al and Berhane et al, there was no significant difference in metabolic syndrome between genders.[^16,18,21^]

Most patients had CD4 counts < 200 at the initiation of ART. The CD4 count of ART-naive group was significantly lower than Zidovudine and Tenofovir groups at the time of recruitment in our study. Patients on the Zidovudine based regimen had a higher prevalence of metabolic syndrome than patients on the Tenofovir based regimen had lower prevalence of metabolic syndrome. Though the differences between the groups were not significant statistically hence are not mentioned. High Blood pressure was most common in the Zidovudine group (*31*.*25%*). High waist circumference was almost equally prevalent in all the 3 groups. Crane et al, through a prospective cohort study of 444 HIV-infected adults without hypertension (at baseline) found that combination therapy with lamivudine and tenofovir as compared with lamivudine and zidovudine was associated with an increased risk of hypertension (*OR, 2*.*3; 95% CI, 1*.*0–5*.*2; P=0*.*046*).[^24^] Similarly, a sub analysis of a prospective cohort study of 527 HIV-infected and 517 HIV-uninfected adults found that prior stavudine exposure was independently associated with hypertension, subsequently leading to more cardiovascular disease (CVD) risk.[^24^]

As per a study done by Sarah J. Masyuko et al., metabolic syndrome was more common amongst HIV-negative than HIV-positive adults, and HIV-negative adults were at greater risk for CVD compared to PLHIV. These data support the integration of routine CVD screening and management into health programs in resource-limited settings, regardless of HIV status.[^25^] Therefore, the PLHIV is as much at risk of developing the metabolic syndrome as much as if not more than the HIV negative patients. Also, the ART drugs do add to the development of cardiovascular diseases independent of the metabolic syndrome. Hence, it is proposed that further studies with larger sample sizes and methodologies providing higher levels of evidence need to be undertaken to study these findings in greater detail. The increased incidence of the metabolic syndrome in the Zidovudine group in comparison with Tenofovir should be taken into consideration, especially amongst populations where Zidovudine based regimens are still used in modern practice. In the metanalysis by Jose Echecopar-Sabogal the protease inhibitors in HIV-infected patients are associated with an increased risk of metabolic syndrome.[^26^] During the collection of data of our study we did not have many patients on protease inhibitors at our center.

In three different cohort studies over a decade, Lucia Taramasso concluded that in PLHIV have a significantly improved metabolic profile compared with previous findings, despite increasing weight and BMI.[^27^]

With the introduction of Dolutegravir based regimens by the government of India, we are facing issues like increased blood sugar levels and obesity.[^1^] Given its possible impact on metabolic syndrome parameters, studies of a similar nature should be repeated to get the most accurate information regarding the current topic in concordance with current treatment regimens.[^10^] Findings of studies such as the present study should be considered to get a holistic outlook on the topic.

## Limitations

A larger sample size would have been more representative of the population with greater statistical power. Information on other environmental factors (diet, activity etc.) was not collected and may be confounders in the above data. Confounding effects of many factors like smoking, alcoholism, oral contraceptive pills, and post-menopausal status could not be eliminated in the present study.

## Conclusion

Males had a significantly higher prevalence of metabolic syndrome than females. The non-nucleoside reverse transcriptase drugs were not associated with metabolic syndrome. Zidovudine had a significantly higher incidence of the metabolic syndrome in comparison with Tenofovir. The Tenofovir based (TLE) regimen was found to be the safest in terms of prevalence of metabolic syndrome (4.2%).

## Data Availability

All data produced in the present work are contained in the manuscript

